# Altered interpersonal distance regulation in autism spectrum disorder

**DOI:** 10.1101/2021.08.06.21261686

**Authors:** Kinga Farkas, Orsolya Pesthy, Anna Guttengéber, Anna Szonja Weigl, András Veres, Anna Szekely, Eszter Komoróczy, Bálint Szuromi, Karolina Janacsek, János M. Réthelyi, Dezső Németh

**Author notes:** **Corresponding author** (KF). **Author contributions** KF, OP, AG, DN contributed to the design and implementation of the research and developed the theoretical formalism/background. KF, AW, OP performed the computations. KF designed the figures. KF, OP, AG, BS, DN wrote the manuscript. OP and AG carried out the experiment. BS and EK contributed to sample preparation. DN, KJ, and RJ supervised the project. All authors provided critical feedback and helped shaping the research, analysis, and manuscript.

## Abstract

Interpersonal distance regulation is an essential element of social communication. Its impairment in autism spectrum disorder (ASD) is widely acknowledged among practitioners, but only a handful of studies reported empirical research in real-life settings focusing mainly on children. Interpersonal distance in adults with ASD and related autonomic functions received less attention. Here, we measured interpersonal distance along with heart rate variability (HRV) in adults with ASD, and tested the modulatory effects of eye-contact and attribution. Twenty-two adults diagnosed with ASD and 21 matched neurotypical controls participated in our study from 2019 October to 2020 February. Our experimental design combined the modified version of the stop distance paradigm with HRV measurement controlling for eye contact between the experimenter and the participant to measure interpersonal distance. Our results showed a greater preferred distance in ASD. These results were altered with eye contact. Moreover, we found lower baseline HRV and reduced HRV reactivity in ASD; however, these autonomic measurements could not predict preferred interpersonal distance. Our study highlights the importance of interpersonal space regulation in ASD and the need for sophisticated experimental designs to grasp the complexity and underlying factors of distance regulation in typical and atypical populations.

## Introduction

Autism spectrum disorder (ASD) is a neurodevelopmental condition characterized by persistent difficulties in social communication and social interaction across multiple contexts, such as abnormal social approach or failure to initiate or respond to social interactions; and restricted, repetitive patterns of behaviour, interests, or activities [1]. At the neural level, cortical [2,3], subcortical [4,5], and autonomic [6,7] neural alterations can be observed, including developmental, structural and functional differences [8–10] in parallel to the pervasive cognitive [11,12], behavioural and physiological disturbances in ASD. However, one of the key components of social behavior, namely interpersonal distance regulation, has received relatively less attention in ASD research (see exceptions: [13–16]) even though its impairment in ASD is widely acknowledged among practitioners. Our study aims to measure the interpersonal distance regulation and a related physiological parameter (heart rate variability, HRV) during this task and test the modulatory effect of two relevant factors in social communication: eye contact and attribution of self or the other, in autism spectrum disorder.

Finding the appropriate social distance can be seen as the first step of physical social interactions. It is widely believed among practitioners that people with ASD keep a greater or abnormal distance [17], violations of personal space occur also more often in ASD in childhood [18]. However, it is challenging to measure this phenomenon experimentally with high ecological validity, and the results are inconsistent. In autistic participants, preference for both closer [15,19–21] or farther distance [16,22] can be found in the literature. Among these studies, only one measured interpersonal distance in adult ASD; they found no difference between study groups [18]. Studies applying electrophysiological or imaging methods usually present video recordings of an approaching individual [14] or use virtual reality displays [23,24] to measure interpersonal distance regulation in ASD. We argue that virtual displays might be useful in training or therapeutic settings, but they cannot take into account several sensory modalities of the scene while measuring the behavioural and physiological reactions in interpersonal interactions. In the present study, we measured the interpersonal distance among adult participants with ASD in an experimental setting with personal presence as close as possible to real-life situations.

Despite its relevance, empirical studies on interpersonal distance regulation of participants with ASD were conducted only in the past few years. The nomenclature of the concept is still not unified. *Personal space* [16,19,22], *social-* or *physical distance* [18], and *interpersonal space* and *-distance* [13,14,16] are all commonly used. When measuring the physical distance between two people in one dimension, we use the term *interpersonal distance*. The Stop Distance Paradigm [25] is the most commonly used method for measuring interpersonal distance regulation. This method is considered an ecological measure of permeability and flexibility of interpersonal space regulation [13]. It can be combined with various factors. Previous studies suggested that the presence of eye contact [19], the role played during the experiment (active approaching or passive indicating) [22], and social interactions as experimental interventions [14,16] might influence interpersonal distance. Most of the studies examined children or adolescents with autism [16,19,22] and found larger preferred interpersonal distance in ASD, except one study [19]. The only study using the Stop Distance Paradigm among adults found no difference between groups in terms of interpersonal distance preferences [18]. Since different results were gained in different conditions, the question arises as to what extent these factors contribute to the outcome behaviour.

Social communication and interactions are tremendously complex processes that can be altered in autism at cognitive, behavioural, and physiological levels. At the highest level, theory of mind difficulties can be observed in ASD [26,27]. To capture this phenomenon during the interpersonal distance measurement in a simplified way, we added a new condition. First, participants had to make a decision based on their own personal preference, next, they were asked to estimate the comfortable distance for the experimenter. This latter situation requires third order mentalization, the mobilisation of theory of mind ability. We called this the attribution dimension. The processing of facial expressions, particularly that of the eye region, is highly relevant in the regulation of social behaviour, including interpersonal distance. Facial emotion processing and emotion recognition is altered in autism [8,28–31]. Constraining eye contact led to an exaggerated increase of amygdala activation, while decreased eye contact was associated with diminished amygdala response to faces in ASD [4,32–34]. In addition, unconsciously avoided eye contact in ASD results in further difficulties of reading socially important signals [35,36]. These results suggest that altered amygdala functioning, including the regulation of eye contact, might have a substantial role in the disturbances of several aspects of social behaviour, such as personal proximity or interpersonal space regulation [25,37,38]. Therefore, in addition to the attribution dimension, eye contact and no eye contact conditions were introduced to investigate the effect of these two factors, which are relevant for social communication.

Physiological response to sensory, social, and emotional stimuli is suggested to be altered in ASD in general, however, the methodology used is highly variable and the results are inconsistent [39]. Since the classic electrophysiological experiment of Hutt et al. showed hyperarousal in children with ASD [40], the majority of studies that measured autonomic regulation (pupillometry, skin conductance, or cardiac measures) found atypical resting-state functions indicating either hyper- or hypoarousal in ASD according to a recent review [41]. Recent studies aimed to overcome these limitations by measuring heart rate variability (HRV): heart rate is affected by both sympathetic and parasympathetic modulatory effects; thus, its variability might be a good marker of autonomic regulation, as higher HRV reflects parasympathetic activity [42]. Furthermore, a study found an association between HRV and cognitive flexibility in healthy individuals [43]. A recent meta-analysis showed that heart rate variability is reduced in ASD: baseline HRV and HRV reactivity during social stress were significantly lower in participants with ASD, but HRV reactivity performing cognitive tasks did not differ [44]. The reduced variability in the heart rate indicates an altered parasympathetic-sympathetic balance in ASD, suggesting the predominance of sympathetic activity and less flexible switching between autonomic states in ASD compared to neurotypicals. For these reasons, we measured interpersonal distance along with heart rate variability to examine their putative alterations and their relationship in ASD.

In this study, our main goal was to establish a comprehensive design to measure interpersonal distance and autonomic functions in ASD. Our first hypothesis was that in adult ASD we observe greater interpersonal distance. Second, we hypothesized that interpersonal distance is modulated by eye contact and attribution. Finally, we aimed to determine the role of autonomic functions in interpersonal distance regulation in ASD, expecting decreased baseline HRV and reduced HRV reactivity during the interpersonal distance task in ASD. It was also hypothesized that autonomic regulation, as characterized by HRV, could predict the preferred interpersonal distance in both study groups.

## Materials and methods

### Participants

In total, 45 adults participated in our research. Two neurotypical participants were excluded due to errors during data collection. The final sample consisted of forty-three participants, 22 were diagnosed with autism spectrum disorder without intellectual disability or language impairment, and 21 were neurotypical controls. The two groups did not differ in age, gender, and education (Table 1). All participants with ASD were diagnosed by trained clinicians, the diagnoses were confirmed with Autism Diagnostic Interview-Revised (ADI-R) and Autism Diagnostic Observation Schedule, IV-module (ADOS-IV.) [45,46]. Twelve participants had one or more comorbid disorders (attention deficit hyperactivity disorder (5), obsessive-compulsive disorder (3), generalized anxiety disorder (2), bipolar disorder (1), depression (1), and schizophrenia (1)). Participants with ASD were recruited from the outpatient unit of the Department of Psychiatry and Psychotherapy, Semmelweis University. Neurotypical control participants were recruited by advertisement.

**Table 1.** Demographics and clinical characteristics.

|  |  | ASD (N=22) |  | NTP (N=21) |  | Statistic | p | effect size |  |
| --- | --- | --- | --- | --- | --- | --- | --- | --- | --- |
| | | N (M/F) | | N (M/F) | | $\chi^2$ | p | | |
| **Gender** |  | 18/4 |  | 14/7 |  | 1.296 | 0.255 |  |  |
|  |  | Mean | SD | Mean | SD | Mann-Whitney U | p | r | 95% CI lower, upper |
| **Age** | (years) | 27.59 | 7.25 | 25.86 | 6.44 | 191.00 | 0.336 | 0.173 | -0.173, 0.481 |
| **Education** | (years) | 15.64 | 3.69 | 16.00 | 3.33 | 248.50 | 0.677 | -0.076 | -0.401, 0.267 |
| **AQ** |  | 31.09 | 6.63 | 15.05 | 5.63 | 18.50 | <0.001 | 0.920 | 0.845, 0.959 |
| **MZQ** |  | 51.68 | 9.53 | 38.29 | 9.30 | 71.50 | <0.001 | 0.690 | 0.462, 0.833 |
| **AAS** | anxious | 22.64 | 5.77 | 16.81 | 6.06 | 114.50 | 0.005 | 0.504 | 0.203, 0.718 |
|  | avoidant | 41.23 | 8.80 | 31.71 | 8.12 | 93.50 | <0.001 | 0.595 | 0.324, 0.776 |
| **ASRS** | Part A | 13.23 | 4.02 | 9.52 | 3.84 | 110.50 | 0.003 | 0.522 | 0.225, 0.730 |
|  | Part B | 26.64 | 9.19 | 16.62 | 5.83 | 88.00 | <0.001 | 0.619 | 0.358, 0.790 |
| **STAI-T** |  | 55.46 | 11.63 | 44.29 | 9.09 | 105.50 | 0.002 | 0.543 | 0.254, 0.743 |
| **ADOS-IV.** |  | 9.96 | 3.35 | - | - | - | - | - | - |
| **ADI-R** |  | 34.82 | 7.74 | - | - | - | - | - | - |
| | | N (Y/N/n.a.) | | N (Y/N/n.a.) | | $\chi^2$ | p | | |
| **Caffeine** | regular | 18/2/2 |  | 15/5/1 |  | 1.558 | 0.212 |  |  |
|  | within 12h | 13/7/2 |  | 13/6/2 |  | 0.051 | 0.821 |  |  |
| **Smoking** |  | 2/18/2 |  | 4/16/1 |  | 0.784 | 0.376 |  |  |
| **Exercise** |  | 18/4/0 |  | 17/3/1 |  | 0.076 | 0.782 |  |  |

ASD: Autism Spectrum Disorder, NTP: Neurotypical Participant, N: sample size, SD: standard deviation, r: rank biserial correlation, 95% CI: 95% confidence interval, M: male, F: female, Y: yes, N: no, n.a.: not available, AQ: Autism-Spectrum Quotient, MZQ: Mentalization Questionnaire, AAS: Adult Attachment Scale, ASRS: Adult ADHD Self-Report Scale, STAI-T: State-Trait Anxiety Inventory - Trait, ADOS-IV: Autism Diagnostic Observation Schedule - IV. module, ADI-R: Autism Diagnostic Interview-Revised

Participants (and legal guardians if applicable) provided written informed consent and did not receive financial compensation for their attendance. The study was conducted in accordance with the Declaration of Helsinki, and it was approved by the Regional and Institutional Committee of Science and Research Ethics, Semmelweis University, Budapest, Hungary (SERKEB No.: 145/2019) from October 2019 to February 2020. The experiment took place at the Laboratory of Brain, Memory and Language Lab, Eötvös Loránd University, Budapest.

### Experimental paradigm - interpersonal distance task

In our study, we measured social distance regulation. Participants underwent an interpersonal distance measurement, a modified version of the stop-distance paradigm [25]. In all conditions, the participant and the experimenter started from the opposite endpoints of the tape measure (five meters) stuck on the ground. They were asked to consciously focus on comfortable social distance overall eight times with the following order. First, (1) participants were approaching actively and were asked to stop where it was still comfortable for them. Next, (2) participants were approaching actively and were asked to stop where they thought it was still comfortable for the experimenter. Then (3) participants stood passively and were asked to stop the experimenter where it was still comfortable for them, finally, (4) participants stood passively and were asked to stop the experimenter where they thought it was still comfortable for the experimenter. Participants repeated this procedure twice, with and without eye contact: either the experimenter was looking at the participant (eye contact condition), or at the papers she was holding (no eye contact condition). The order of these two conditions was randomised across participants (Fig 1). During the statistical analysis active and passive conditions were pooled (averaged) together.

**Fig 1.**
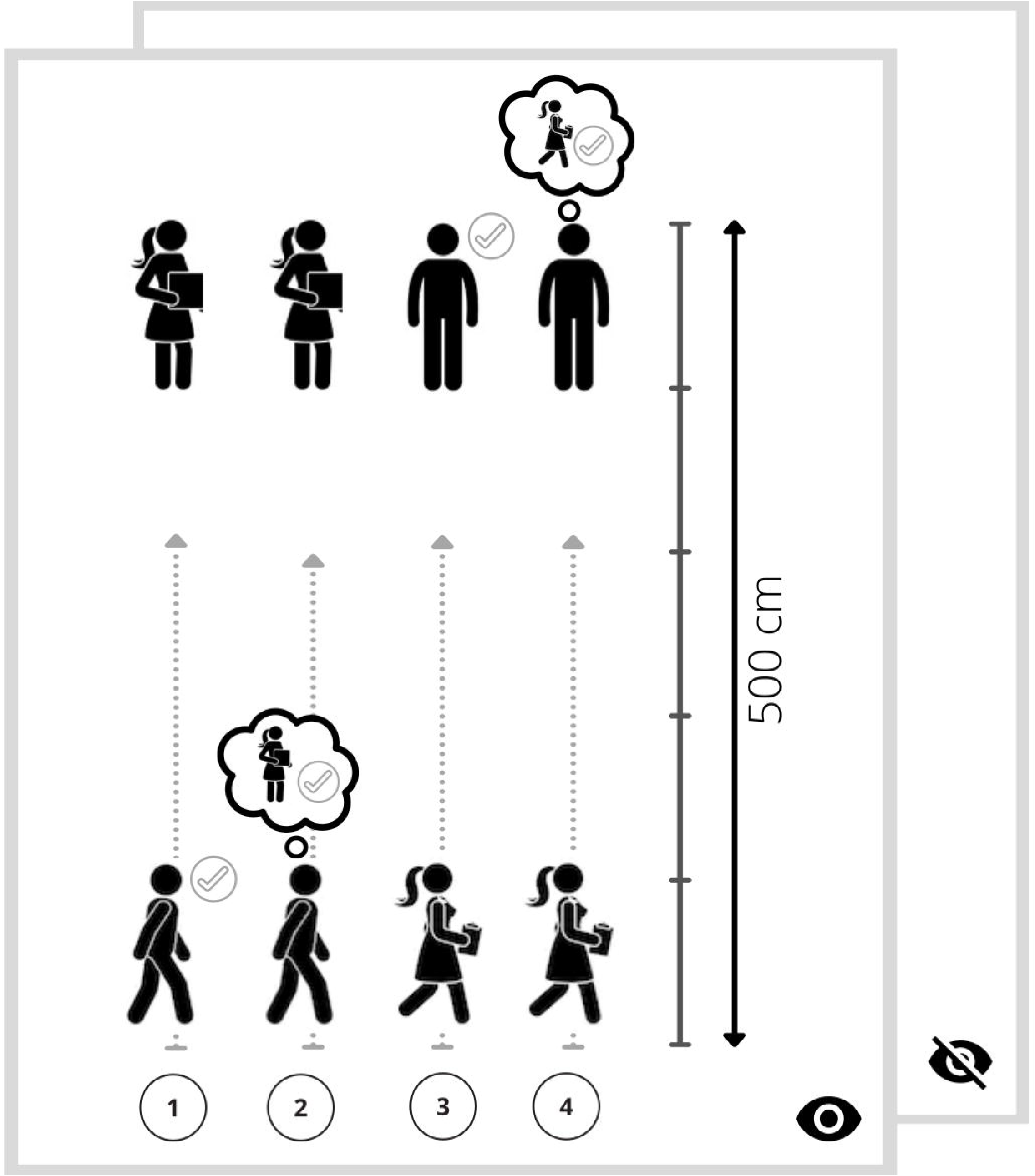
Experimental setting. The modified version of the stop distance Paradigm. First, (1) participants were approaching actively and were asked to stop where it was still comfortable for them. Next (2) participants were approaching actively and were asked to stop where they thought it was still comfortable for the experimenter. Then (3) participants stood passively and were asked to stop the experimenter where it was still comfortable for them; finally (4) participants stood passively and were asked to stop the experimenter where they thought it was still comfortable for the experimenter. Participants repeated this procedure twice with and without eye contact; the order of the latter two conditions was randomised across participants.

### Heart rate monitor

A wearable device Polar H10 was placed on participants’ chests which recorded heart rate (HR) during the whole experiment. We measured cardiac interbeat intervals (RR intervals) using Polar H10 heart rate monitor chest strap (Polar Electro Oy, Kempele, Finland) [47], which has been shown to be a valid device to measure RR interval signals [48]. The HR monitor was connected to a Samsung Galaxy Tablet via Bluetooth. We used the Elite HRV application to export the recorded RR intervals as .txt files. We measured heart rate variability (HRV) at two different conditions for a duration of 60 seconds: 1) at baseline and 2) during the intentional interpersonal distance task (1 minute after starting the distance task) using the Root Mean Square of Successive RR interval Differences (RMSSD) method [49]. Additionally, we calculated RMSSD at the preceding ten-second time window of trigger points set by researchers. These triggers corresponded to the time when participants arrived at their final location of each condition.

### Distance measuring: Obimon Prox

In order to synchronize the distance data with the HRV data, both the experimenter and the participant wore a distance measuring device. The Obimon Prox [50] measures the distance and the relative orientation between two wearable devices in real-time. The devices use Ultra Wide Band (UWB) technology and the Symmetrical Double Sided Two-Way Ranging (SDS-TWR) method [51] to both determine the distance between each other by emitting very short and low power radio transmissions and measure the so-called time-of-flight (ToF) with very high precision between transmission and reception. The resolution of the measurement is in the range of a few centimetres, while the absolute precision is approximately 10 centimetres. The relative orientation is defined as the difference between the angles between the two devices taking the Earth’s magnetic field as a reference. For increased precision, the device uses sensor fusion involving magnetometer, accelerometer, and gyroscope sensors. The results of the measurements are collected over Bluetooth LE wireless technology to a laptop computer and evaluated in real-time.

### Procedure

Participants wore a Polar H10 and Obimon Prox device during the whole experiment. They placed the wearable device on themselves before the experiment started, then they waited five minutes while calibrating and registering 60 seconds of resting heart rate and HRV.

Next, participants completed the interpersonal distance task. Then, after a short break, they completed a computerized neurocognitive test battery - measuring working memory, executive functions, attention, inhibition, implicit learning, faux pas. These results are not reported in this paper. Finally, they completed computerized versions of self-report questionnaires (AQ: Autism-Spectrum Quotient, MZQ: Mentalization Questionnaire, AAS: Adult Attachment Scale, ASRS: Adult ADHD Self-Report Scale, STAI-T: State-Trait Anxiety Inventory – Trait; see Table 1 and Supporting information, S1 File).

To avoid the sensory over-reactivity effect, experimenters did not wear any jewellery, perfume, and had been asked not to eat spicy food before the experiment, they wore simple, casual, non-coloured clothes (jeans and black T-shirt). The room was curtained and artificially evenly lit.

### Data preprocessing and analysis

Preparation of HRV data was carried out using Python 3.7 with NumPy 1.20.1 [52], pandas 1.2.3 [53], and SciPy 1.6.1 [54] data processing packages. Since the samples were measured at a different rate for the Polar H10 (one sample per second) and the Obimon prox (one sample per milliseconds) devices, we resampled the Obimon data by taking the median for each second. Missing data were dropped from the analysis. To synchronize HRV with the proximity data we needed to obtain the timestamps for each file containing the RR intervals. The first timestamp was obtained from the name of the file which indicated the start time of the recording. Since the exported files only contained the RR intervals without a timestamp for each sample, the interval values themselves were used to create the time elapsed since the first sample. As RR intervals annotate the time between two successive heartbeats, it was possible to append the value of the RR interval to the time of the previous sample. After obtaining the timestamps, data points were replaced with the median if they indicated RR of 1200 milliseconds (ms) or above, or if their absolute Z score was higher than 2. Triggers added to the distance data (see Distance measuring) were adjusted manually if needed. HRV was estimated as the root mean square of successive RR interval differences (RMSSD) since this measurement is relatively resistant to by-products caused by breathing [55], and can be obtained for a shorter (10 seconds) period of time [56]. Calculations were done by the following formula (1):

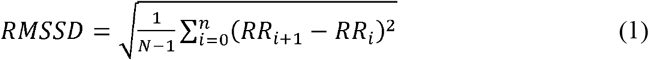

Baseline HR and HRV were measured and calculated for 60 seconds (s) at baseline, reactive HR and HRV were measured during interpersonal distance task one minute after starting the explicit paradigm (from +60 s to +120 s).

Furthermore, we calculated RMSSD around time points where interpersonal distance data were reported. To calculate RMSSD for each explicit condition, eight local minimums of the distance data were determined from data recorded by Obimon Prox. These eight time points indicate the shortest distances between the participant and the experimenter, corresponding to the time point when the reported distance was reached. RMSSD was calculated for an interval starting 10 seconds prior to reaching the reported distance.

### Statistical analysis

Statistical analysis was accomplished using R Version 3.6.3 [57], RStudio Version 1.2.1335 [58], and JASP Version 0.14.0.0 [59]. First, to measure if the two study groups do differ regarding age, gender, education, caffeine intake, smoking, exercise and scores on questionnaires, we conducted nonparametric Mann-Whitney U (Wilcoxon rank-sum) tests and a Chi-square test. To measure the effect of different conditions and study groups on the distance and HRV data, mixed-design ANOVA tests were applied, while in the case of significant interaction effects post hoc tests with Bonferroni correction were used. As there was already a baseline difference in HRV between the two groups, HRV values were standardised in the interpersonal conditions for the further comparisons. Associations between distance, HRV, and scores of psychometric questionnaires and diagnostic tests were analysed with Spearman’s rank-order correlations. Analyses were performed and visualizations were created with R-packages *dplyr* [60], *ggplot2*[61], *psych* [62], *gridExtra* [63], *ggpubr* [64], *readxl* [65], *corrplot* [66], *Hmisc* [67], *varian* [68].

## Results

### Is preferred interpersonal distance different in ASDã

Participants with ASD set significantly greater distances than NTP participants on a group level. For the descriptive statistics of all conditions see supplementary material (Table S1). To test if the different conditions had different modulatory effects in the two study groups, we used two-way mixed-design ANOVA on the interpersonal distance as dependent variable, where the between-subject factor was the Group (ASD/NTP), within-subject factor Condition (Eye contact, Attribution). The Group main effect *F*(1,41)=8.999, *p*=.005, *η*^*2*^_*p*_ =0.180 was significant, participants with ASD preferred larger distances in general (mean difference=35.980, 95% CI [11.757, 60.203]) (Fig 2). According to the post-hoc power analysis, the group difference was detected with 98% power. Levene’s test showed that the variances were equal.

**Fig 2.**
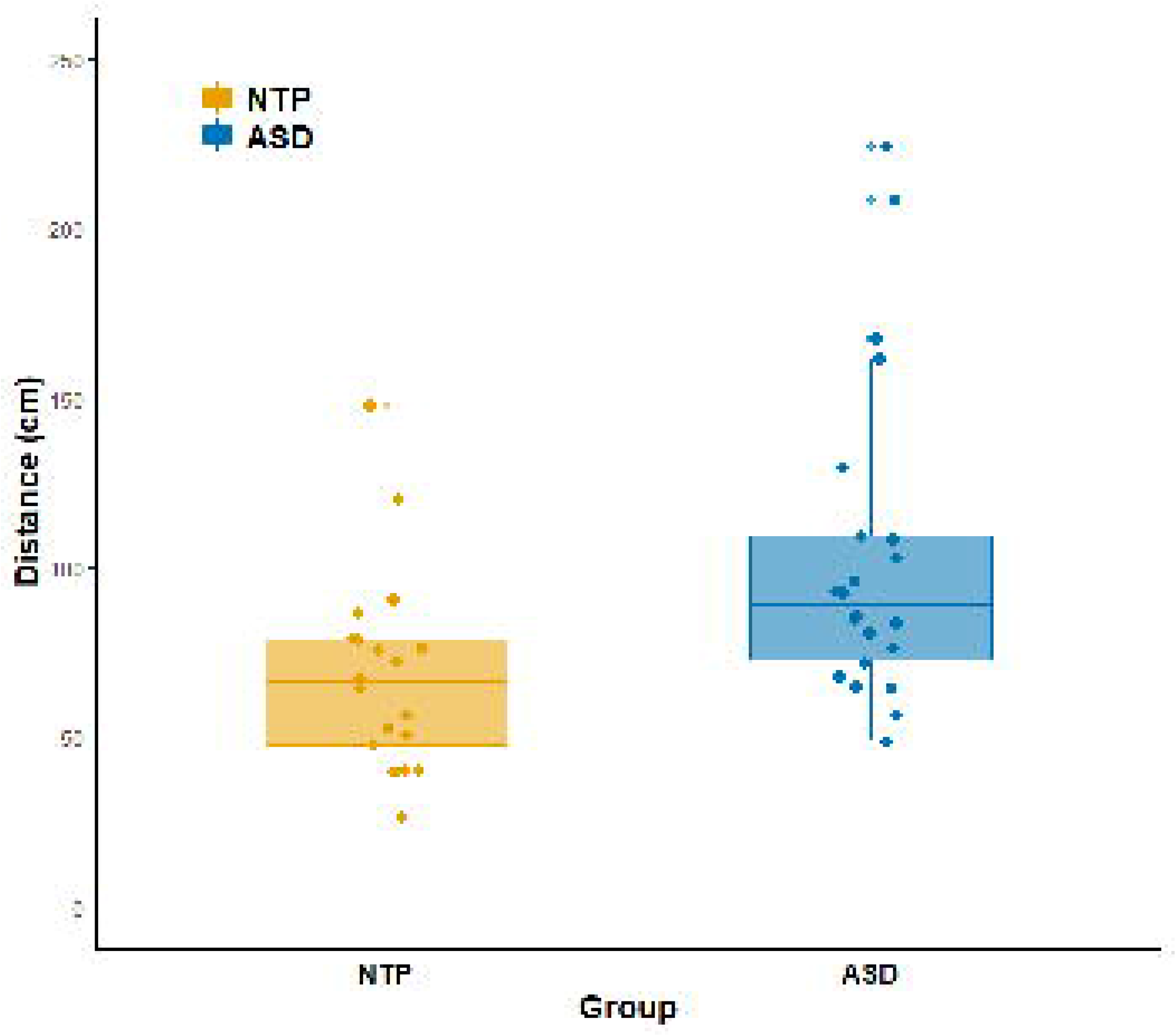
Interpersonal distance in cm. Dots represent the mean of distance data of eight conditions for each individual. The top and the bottom of the box show the upper (Q3) and lower (Q1) quartiles, the line dividing the box represents the median, and notches show a 95% confidence interval around the median. Asterix indicates significant group difference. Orange: neurotypical participants, blue: participants with ASD.

### Does eye contact or attribution affect interpersonal distance?

The main effect of the conditions of eye contact, or attribution resulted in the following. Eye-contact (*M*_eye_(*SD*) = 88.756 (47.616), *M*_no_eye_(*SD*) = 83.442 (40.116), *F*(1,41)_eye_cont_ =3.005, *p*=.091,*η* ^*2*^_*p*_ =0.068, 95% CI [−0.859, 11.267]) showed a trend, and attribution (*M*_self_(*SD*) = 84.913 (53.165), *M*_other_(*SD*) = 87.285 (37.378), *F*(1,41)_attrib_=0.248, *p*=.621, *η* ^*2*^_*p*_ =0.006, 95% CI [−12.691, 7.671]) did not have a significant main effect, indicating that participants attributed similar comfortable personal distance to the experimenter as to themselves (Fig 4 a-b).

In the further analysis, we tested if the conditions influence each other’s effect. We found a trend in Eye-contact × Attribution interaction (*F*(1,41)=3.011, *p*=.090, *η* ^*2*^_*p*_ =0.068) but no significant Group × Condition interaction. Post-hoc test revealed that there was the biggest difference between NTP and ASD participants when they had to maintain eye contact and they had to set the comfortable distance for themselves (*t*=3.426, *p=*.030, 95% CI [2.248, 90.457]) after adjusting p-values for multiple comparisons by Bonferroni-correction in post hoc tests.

### Are heart rate and heart rate variability altered in ASD?

In general, participants with ASD had a slightly higher heart rate (*M*_*baseline*_*=*90.65, *SD*=12.95; *M*_*interpersonal*_=96.66, *SD*=12.49) than NTP participants (*M*_*baseline*_*=*87.06, *SD*=15.74; *M*_*interpersonal*_=91.68, *SD*=15.08) (Figure 3), however, these differences were not significant. Measuring the effect of the conditions we used mixed-design ANCOVA, with the between-subject variable of Group (ASD/NTP), and within-subject variable Condition (baseline/interpersonal). As caffeine intake could influence the heart rate, we included actual caffeine use as a covariate, but it did not change the results. The Group main effect was not statistically significant (*F(1,35)*=0.875, p=.356, *η* ^*2*^_*p*_ =0.024), the main effect of Conditions was significant (*F*(1,35)=38.068, *p*<.001, *η* ^*2*^ =0.521), but the Condition × Group interaction was not (*F*(1,35)=0.647, *p*=.427, *η* ^*2*^_*p*_ =0.018). It means, that in both groups we measured the highest HR during the intentional interpersonal distance task, and it was significantly higher than baseline (*t*_ASD_=5.866, *p*<.001, 95% CI [2.554, 9.454], *t*_NTP_=3.846, *p*=.003, 95% CI [1.260, 7.977]) according to the post-hoc tests, where *p* values were adjusted by using Bonferroni-correction. Levene’s test showed that the variances were equal (Figure 3a).

**Fig 3.**
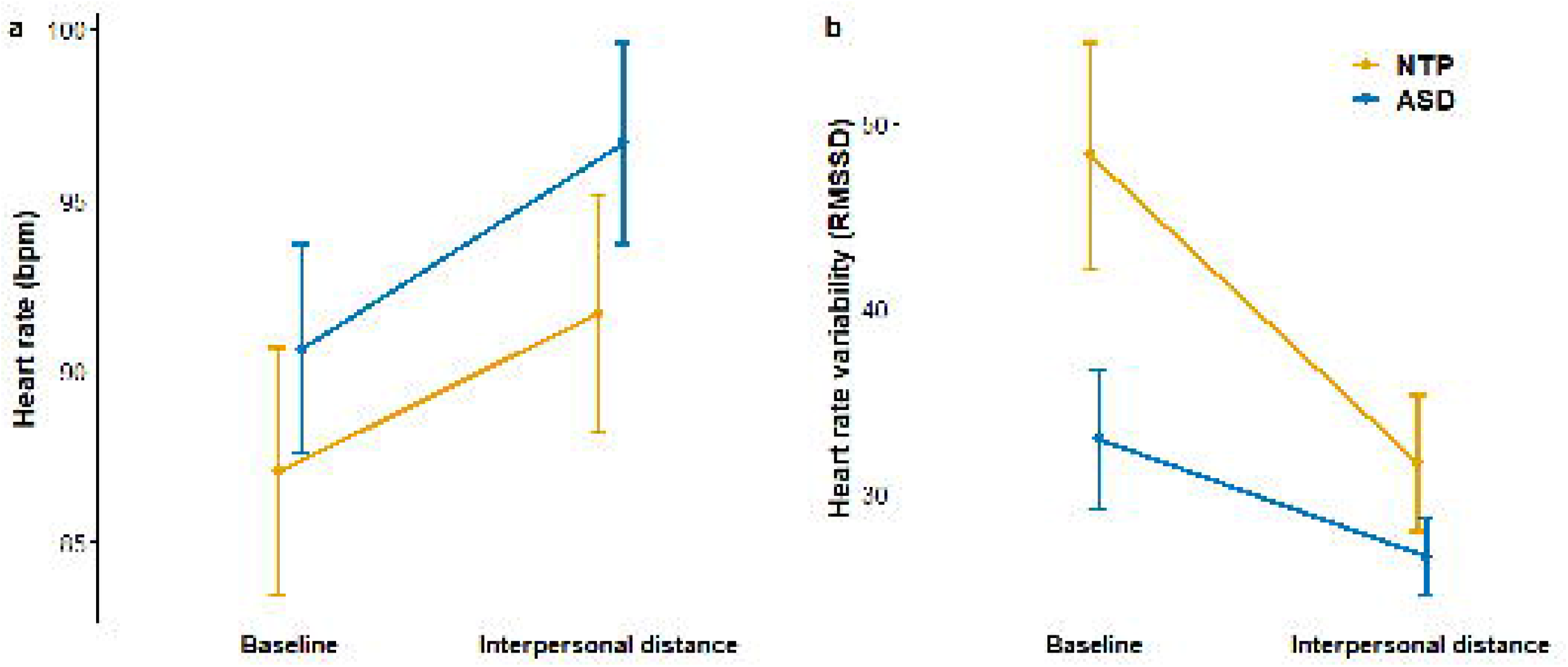
Heart rate and heart rate variability. Panel A: Baseline and reactive (interpersonal conditions) heart rate in beat per minute (bpm). Panel B: Baseline and reactive (interpersonal conditions) heart rate variability (RMSSD). Error bars: standard error of the mean. Asterix indicates significant group difference. Orange line: neurotypical participants, blue line: participants with ASD.

Heart rate variability (HRV) was higher in the NTP group (*M*_*baseline*_*=*48.26, *SD*=26.68; *M*_*interpersonal*_=31.60, *SD*=16.11) than in ASD (*M*_*baseline*_*=*32.90, *SD*=15.97; *M*_*interpersonal*_=26.57, *SD*=8.79). Again, measuring the different effects of the conditions we used mixed-design ANOVA, with the same within- and between-subject variables described above. The group main effect (*F*(1,35)=3.470, *p*=.071, *η* ^*2*^_*p*_ =0.090) showed a trend. The condition main effect (*F*(1,35)=22.744, *p*<.001, *η* ^*2*^_*p*_ =0.394) and Group × Condition interaction (*F*(1,35)=4.598, *p*=.039, *η* ^*2*^_*p*_ =0.116) were significant. Post-hoc test showed significant difference between the baseline and interpersonal condition (*t*=4.769, *p*<.001, 95% CI [6.600, 16.383]), but this difference originates from the significant difference within the NTP group (*t*_NTP_=4.956, *p*<.001, 95% CI [7.258, 26.059]), whereas HRV did not differ significantly in ASD group between the two conditions (*t*_ASD_=1.831, *p=*.276, 95% CI [−3.333, 15.982]) (Figure 3b). The more pronounced difference between baseline HRV and HRV during the task (reflective HRV) suggests a greater reactive capacity of autonomic regulation in NTPs.

### Does eye contact or attribution affect interpersonal heart rate variabilityã

HRV during the interpersonal distance task (measured before the time point reported distance was reached) was numerically higher in the NTP group, but the difference was not significant between groups. For descriptive statistics see Supplementary material (Table S2). As the baseline HRV was higher in NTP group, we used standardized HRV here. When we tested the effect of the conditions and their interactions, neither the main effect of eye contact or attribution, and their interaction with the group, nor the group effect (*F*(1,32)=0.0002, *p*=.988) were significant using repeated-measures ANOVA. (Fig 4 c-d). Autonomic functioning might be influenced by smoking, exercise, regular caffeine consumption, or the actual caffeine intake before the experiment. There was no difference between groups (see Table 1), however, including these variables as covariates did not change the results.

**Fig 4.**
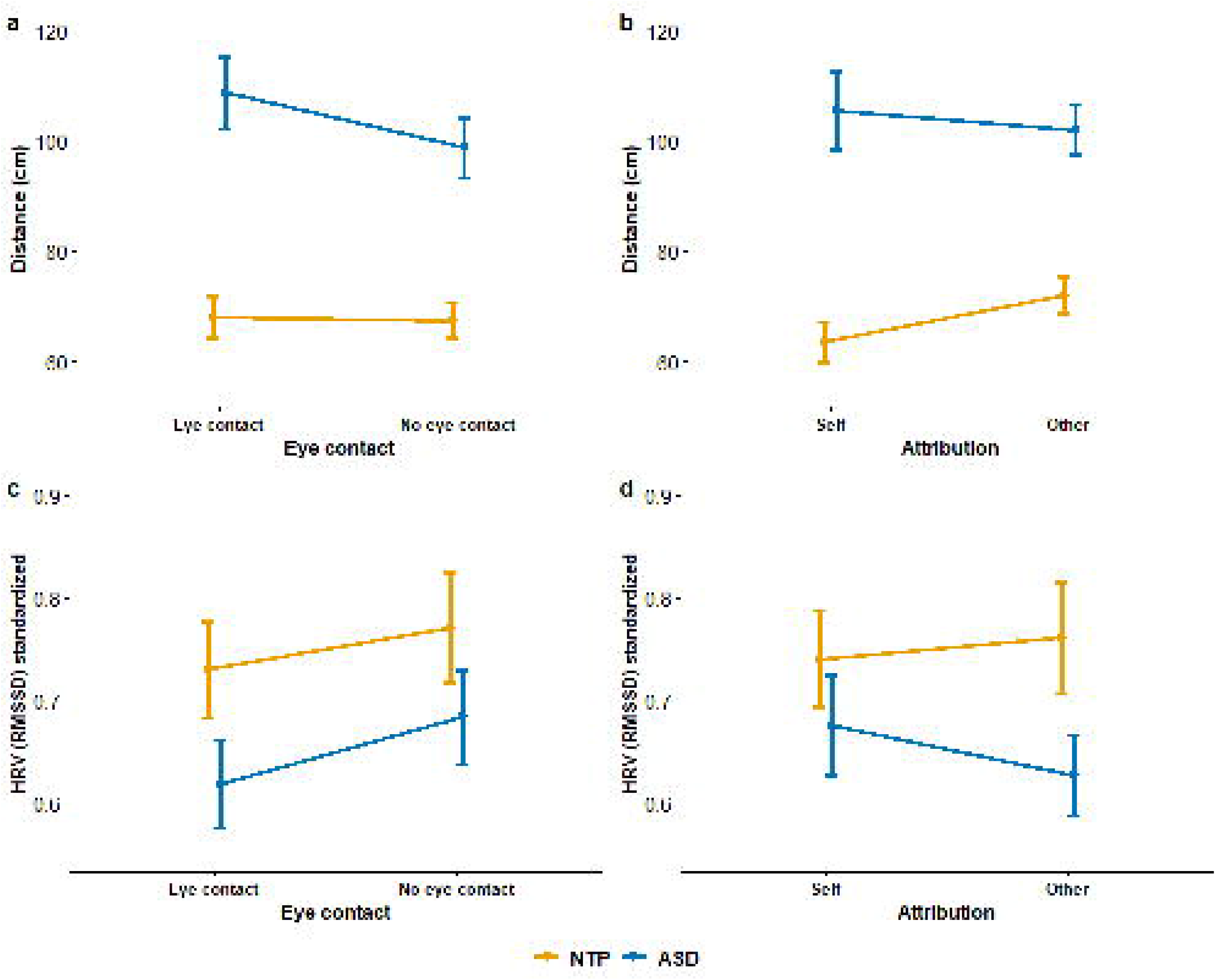
Interpersonal distance and HRV differences in different conditions. Panel a-b Interpersonal distance in cm. Panel a: With or without eye contact, Panel b: Attribution to self or the other. Panel c-d Heart rate variability in explicit conditions at a reported distance. Panel c: With or without eye contact, Panel d: Attribution to self or the other. Error bars: standard error of the mean. Asterix indicates significant group difference. Orange line: neurotypical participants, blue line: participants with ASD.

The interpersonal distance (Panels a-b) and heart rate variability data measured by the RMSSD method (Panels c-d) are presented in Fig 4 to introduce their characteristics in different conditions in the two study groups.

### Is there any correlation between HRV, distance, and psychometric data?

#### Exploratory analysis

Correlation analysis was highly exploratory, due to the small sample size, but it might be suitable for further hypothesis generation. We found a relatively high consistency of distance and HRV data (Fig 5), suggesting both the measured distance and HRV data indicate reliably a similar construct. Intentional interpersonal distance and baseline HRV data show a weak negative correlation in NTP participants, suggesting that higher heart rate variability fosters prosocial activity and neurotypical participants with greater autonomic regulation could approach the experimenter more (Fig 5, upper triangle). In the case of participants with ASD, however, we found less consistent association between distance and HRV results (Fig 5, lower triangle). Correlation between the mean interpersonal distance and HRV during the interpersonal distance task, was not significant, however, it tend to point in different directions in the two groups (Fig 6).

**Fig 5.**
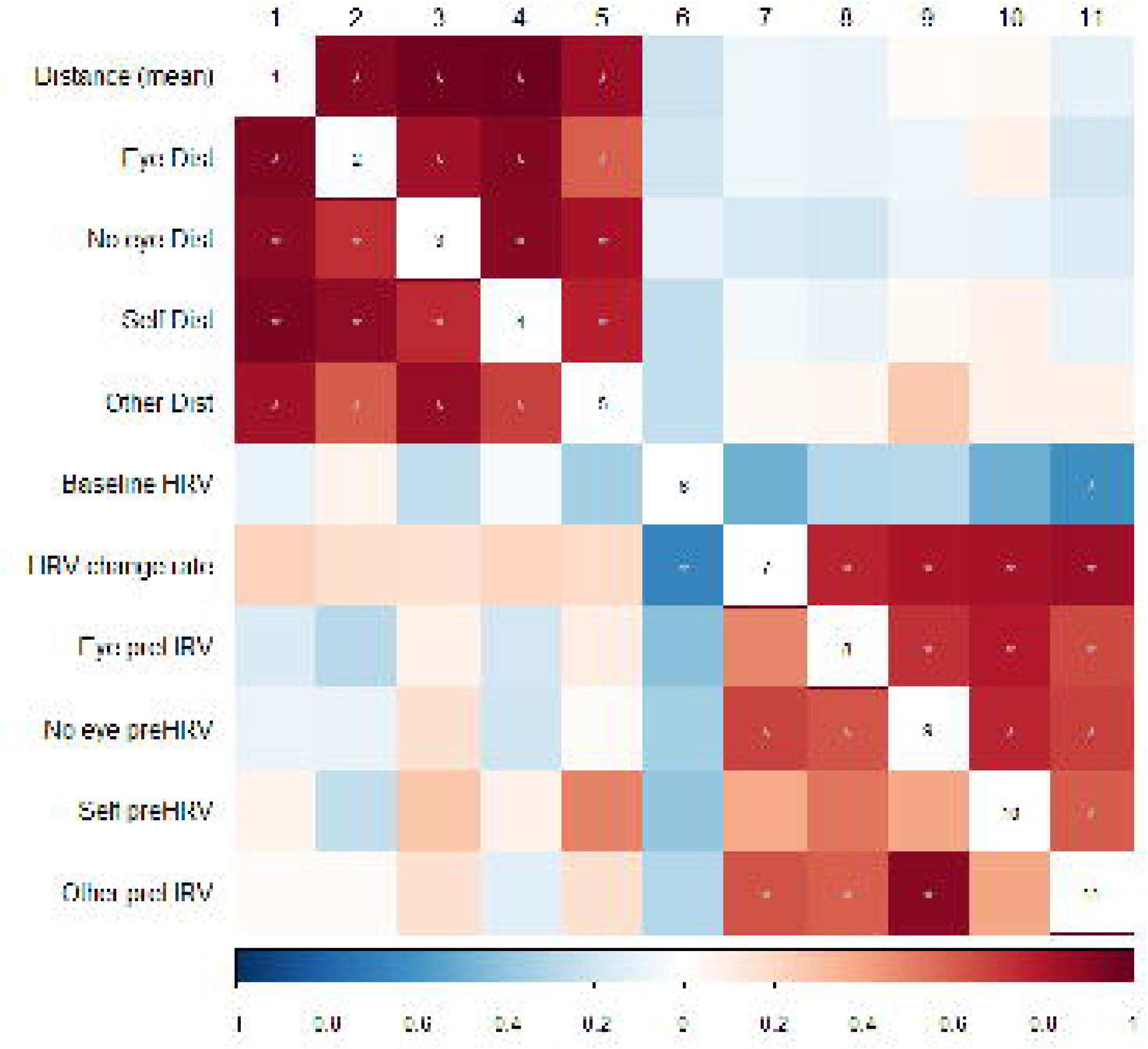
Correlations between interpersonal distance and heart rate variability at the baseline and during the intentional interpersonal distance conditions. Dist = distance, HRV = heart rate variability, preHRV = 10s RMSSD, Eye = eye contact, No eye = no eye contact, Active = active moving, Passive = standing, Self = attribution to self, Other = attribution to the other conditions. Upper triangle: neurotypical participants, lower triangle: participants with ASD. Warm colors refer to positive, cold colors refer to negative Spearman rank correlation *rho* values, grey asterisk marks the significant *p* values after (fdr) correcting for multiple comparisons.

**Fig 6.**
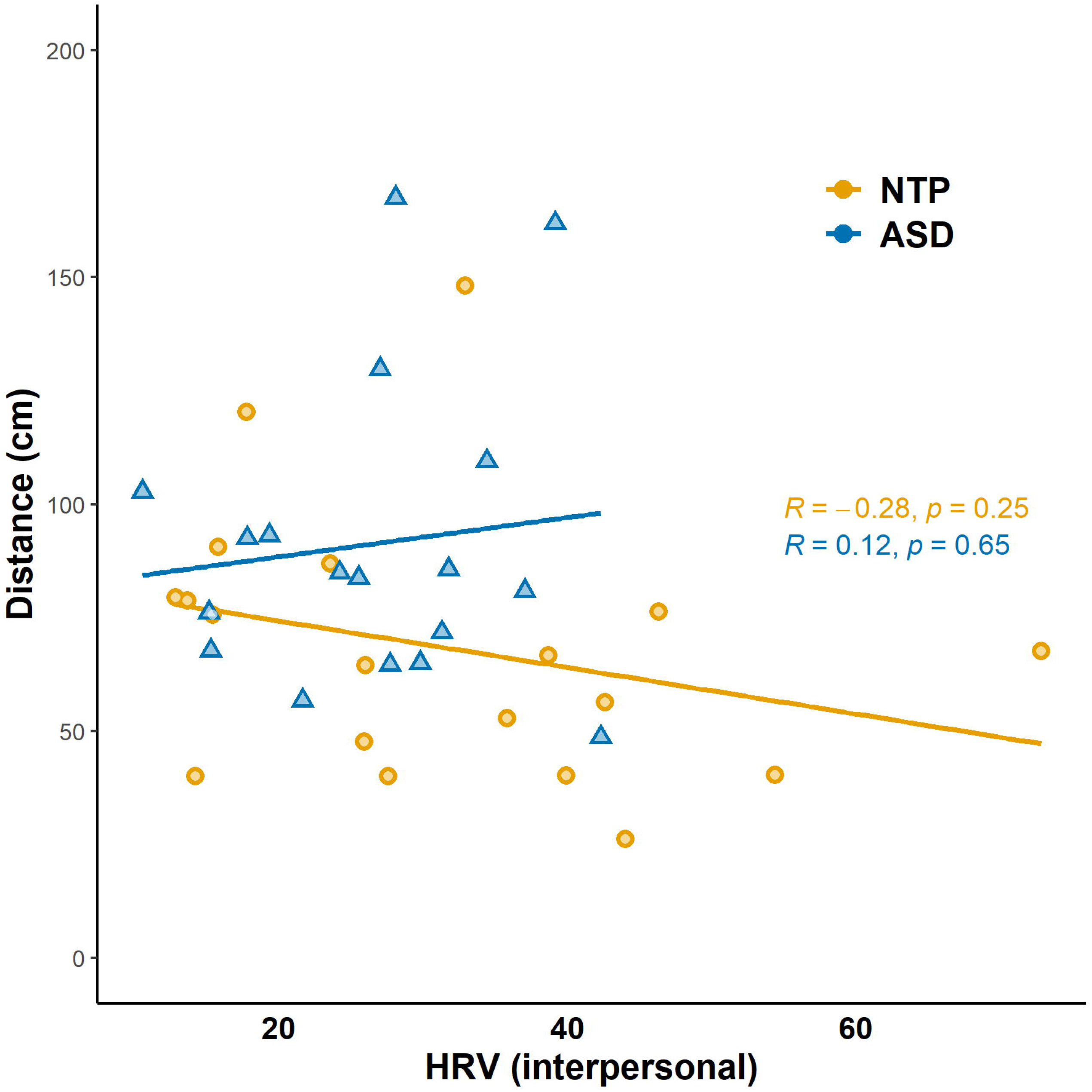
Correlation between mean distance (in cms) and HRV (60 s) during interpersonal distance task in the two groups. Orange line: neurotypical participants, blue line: participants with ASD

Results of psychometric questionnaires showed weak or no association with distance and HRV results (Fig 7); however, the association with psychometric questionnaires in ASD showed a different pattern than in NTP. High trait anxiety level, poor mentalization, and attachment were weakly associated with greater interpersonal distance ASD (Fig 7, lower triangle, first column), but neither of these correlations remained significant after correction for multiple comparisons, only HRV at baseline and during the interpersonal condition, AQ and mentalization scores were correlated in ASD group.

**Fig 7.**
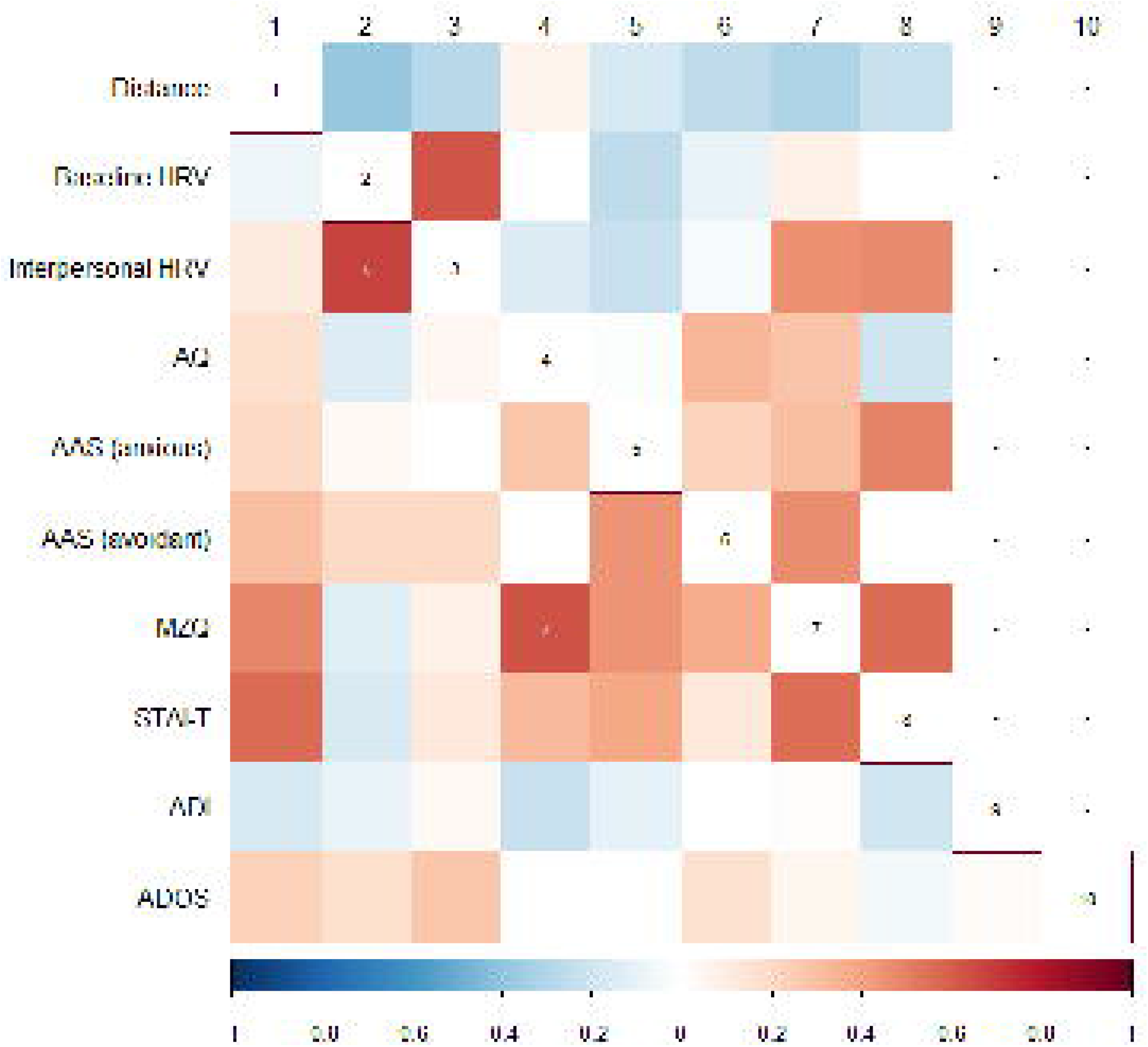
Correlations between interpersonal distance, heart rate variability at the baseline and during the interpersonal distance conditions, and psychometric data. HRV = heart rate variability, AQ = Autism-spectrum Quotient, AAS = Adult Attachment Scale, MZQ = Mentalization Questionnaire, STAI-T = State-Trait Anxiety Inventory, Trait, ADI = Autism Diagnostic Interview-Revised, ADOS = Autism Diagnostic Observation Schedule. Upper triangle: neurotypical participants, lower triangle: participants with ASD. Warm colors refer to positive, cold colors refer to negative Spearman rank correlation *rho* values, grey asterisk marks the significant *p* values after (fdr) correcting for multiple comparisons.

## Discussion

Our study aimed to investigate interpersonal distance regulation and the underlying autonomic response regulation in autism spectrum disorder. To this end, we introduced a methodology combining interpersonal distance measurement and physiological parameter registration in an interpersonal experimental setting in groups of adult participants with ASD and their matched neurotypical controls. We found increased interpersonal distance in ASD. The difference was the most pronounced when the experimenter maintained eye contact and requested to determine the comfortable distance for themselves during the interpersonal distance task. Moreover, we found decreased baseline heart rate variability and decreased HRV reactivity in ASD.

Interpersonal distance was measured using a modified version of the stop distance paradigm to assess how far participants prefer to stand from another person and whether there is a difference between ASD and NTP group in this respect. Participants were directly instructed to define a still comfortable distance from the experimenter. Usually, in a stop distance paradigm, the participant and the experimenter are facing each other at the endpoints of a 300 to 600 cm long line along which the participants set their preferred interpersonal distance. In our experiment, we have chosen 500 cm as the initial distance, which includes all four distance zones of interpersonal space (intimate, personal, social and public) according to Hall’s proxemic rules [69]. During this task, participants set distances on average within the personal space (far zone ∼75 - 120 cm, close zone ∼45 - 75 cm, the zones between the intimate distance and personal distance). However, as expected, participants with ASD set significantly greater distances than NTPs: around the far end of the personal space, or even farther. The social space (the zone between personal and social distance 120 - 370 cm) is reserved for strangers or new acquaintances [69]. We can speculate that this display can change the non-verbal message conveyed by a set distance in real-life interpersonal situations.

Prior to our experiment, five studies examined interpersonal space regulation in ASD, four of which studied children or adolescent populations. These findings uniformly suggest that interpersonal space regulation is altered in ASD in childhood, autistic children preferred significantly larger interpersonal distance than neurotypical control participants [16,22]. A study examining adolescents with ASD also concluded that their space regulation was altered. Interestingly, this conclusion was derived from opposing results: adolescents with ASD preferred shorter interpersonal distance than neurotypical controls [19]. Although, in this study from Japan, neurotypical participants preferred longer personal distances (cca. 130 - 150 cm depending on condition) than in other cohorts, and the distances preferred by ASD participants were comparable to our and Kennedy et al.’s results, the only adult study observed no differences between participants with ASD and neurotypical controls [18] (cca. 70 – 100 cm). This raises the possibility of cultural differences in social rules and customs, including personal space arrangement. Our results suggested that adult individuals (a homogenous white Caucasian, Central European sample) with ASD prefer greater interpersonal distance than neurotypicals.

Several factors can influence the interpersonal distance between the experimenter and the participants [13,19,22]. Eye contact has been shown to affect the preferred interpersonal distance of ASD and neurotypical adolescents: in eye contact conditions when participants held passive roles, they preferred larger interpersonal distance, regardless of which study group they belonged to, and this effect did not emerge when holding active roles [19]. In our study we found a weak modulatory effect of eye contact, participants with ASD tend to set slightly greater distances in conditions with eye contact. We can speculate that eye contact might cause distress and evoke avoidance in ASD, instead of an appetitive or comforting positive reinforcement, in line with the “eye avoidance” hypothesis [33,34]. Future studies directly examining this phenomenon and testing whether and how eye contact plays a role, seem warranted.

Reciprocal social interactions are impaired in ASD, leading to a weaker adaptation to another person’s perspective. Therefore, we also introduced a condition that requires higher order mentalization. We tested what the participant thought was a comfortable distance for the experimenter. Considering the theory of mind difficulties in ASD, we hypothesized that participants with ASD will show less difference along the attribution dimension than NTPs. We found no difference in ASD accordingly, although there was no significant difference between the self and other attribution conditions in the NTP group either. It is interesting, however, that we measured the biggest distance in ASD and biggest difference between NTP and ASD participants when they had to maintain eye contact and they had to set a comfortable distance for themselves. These results might suggest that participants with ASD are capable of modifying their behaviour to some extent according to others’ aspect, in contrast with previous results [70,71], but to test this hypothesis further studies are needed.

An invention in our experimental design is that we combined interpersonal distance measurement with heart rate registration. In social behaviour parasympathetic regulation, the flexibility of vagal tone plays an important role according to Porges’ polyvagal theory [72– 74]. Higher resting HRV was found to be associated with cooperative behaviour, using less disengagement and more socially adaptive emotion regulation strategies among healthy adults [75,76]. Variables influenced by parasympathetic regulation (e.g. respiratory sinus arrhythmia) are related to emotion recognition and symptom severity in ASD [77]. In line with the results of previous studies corroborating altered autonomic nervous system functioning in ASD [41,44,78], we found reduced baseline heart rate variability in participants with ASD spectrum disorder. The average heart rate (reflecting sympathetic activity) was slightly higher in the ASD group than in neurotypicals, but this difference was not significant.

In social situations, not just the baseline but also the regulatory capacity must be taken into account. Previous studies showed that the HRV decrease induced by participating in a social situation was lower, assuming a decreased regulatory capacity in the ASD group [44]. We used the RMSSD method to measure HRV in order to capture the parasympathetic regulation rather than sympathetic arousal [49]. Psychotropic medication, caffeine intake, smoking, and exercising habits might influence autonomic regulation [79–82], but study groups in this study did not differ in actual or regular caffeine intake, smoking and exercising habits (Table 1). There were no significant differences between participants who took medication and those who did not, consequently, we argue that the results are attributable to autism. We found a significant HRV decrease in interpersonal setting compared to the baseline in NTP, but not in ASD group, which confirms previous research findings of decreased regulatory capacity of participants with ASD in social situations.

Additionally, we calculated HRV during the social distance regulation task in 10 s time periods, applying ultra-short-term analysis [56] exactly at the time point when participants arrived at the reported distance in order to take a closer look at the relationship of interpersonal distance and autonomic regulation. To test experimentally the more nuanced aspects of interpersonal distance regulation, we assessed the modulatory effect of eye contact and attribution. We did not find a significant difference between study conditions regarding the 10 s HRV metrics, and results did not directly support our hypothesis that HRV predicts the interpersonal distance. Nevertheless, we can speculate that due to the inherently lower baseline HRV in ASD, the diminished capacity of reactive decrement might have prevented the further fine-tuning during the interpersonal task. Reduced regulatory capacity, combined with elevated amygdala reactivity could lead to early exhaustion even during minimal social interaction. This can raise the possibility that the larger interpersonal distance is the consequence of the early exhaustion of regulatory capacity, and by keeping the distance they might avoid a more severe autonomic disturbance in social situations. To further test this hypothesis in real-life situations, applying widely available wearable devices might be useful. This experience gained could later be used, for example, to develop biofeedback tools for social communication training for people with autism spectrum disorder.

### Limitations and further directions

Despite the most careful planning, every study has its limitations. In this study, we examined adult participants with ASD trying to fill a gap in this research area and measure interpersonal distance and autonomic regulation simultaneously. We recruited participants with average or above-average intellectual abilities, which increases the likelihood of adaptive skill acquisition. To overcome this limitation inclusion of a broader spectrum of autistic participants is needed in future studies.

We found greater interpersonal distance in ASD measured by the modified version of the stop distance paradigm, but there was no difference between study groups regarding heart rate variability during that part of the experiment. In subsequent research, HRV differences should be measured at fixed distances as well, e.g. closer than the comfortable range for neurotypicals, farther than the comfortable range for participants with ASD, and at intermediate distances. Subjective rating of the level of comfort (both by participants and by the experimenter) might help to gain a better insight of how correctly the experimenter’s perspective can be estimated by the participants.

Unfortunately, we have had to finish the data collection due to the beginning of the COVID-pandemic. As the introduced social distancing measures deeply affect our senses of interpersonal distance, we cannot continue with the original study design. We plan to investigate the effect of this unforeseen but global intervention by repeating our experiment when the pandemic subsides, and the social aspects of the situation are consolidated. Additional conditions with and without wearing face masks might be considered, too. These subsequent studies will be able to show us whether autistic people have been affected differently than neurotypicals by social distancing measures.

## Conclusion

Interpersonal distance regulation is a relevant nonverbal part of social communication. It reflects the individual needs for personal space and the ability to read others’ intentions. Together with other biomarkers of autonomic functions, this might express how demanding a simple social interaction can be for people with ASD. In this study, we introduced a new experimental design to measure these factors together, in a basic social interaction setting. Participants with ASD preferred larger interpersonal distance. The difference was most pronounced when they had to approach the experimenter actively with maintained eye contact. Participants with ASD had lower baseline heart rate variability and decreased heart rate variability reactivity than neurotypicals; however, their HRV and its changes in different conditions during the social distance task did not differ significantly. This raises the possibility that regulatory capacities were exhausted sooner, at a farther distance in this group, as part of the compensatory avoidant behaviour in ASD. We believe that applying this experimental design could also be beneficial in studying other psychiatric conditions, such as borderline personality disorder, social phobia, or psychosis. These results could further expand our understanding of interpersonal distance regulation in autism spectrum disorder.

## Data Availability

The data that support the findings of this study are available on request from the corresponding author.

## Acknowledgment

This research was supported by the National Brain Research Program (project 2017-1.2.1-NKP-2017-00002, PI: D. N.); the Hungarian Scientific Research Fund (OTKA K 128016, PI: D. N., OTKA PD 124148, PI: K. J.); the János Bolyai Research Scholarship of the Hungarian Academy of Sciences (to K. J.); the New National Excellence Program of the Ministry for Innovation and Technology (ÚNKP-19-2-I-ELTE-332, A. G.); the IDEXLYON Fellowship of the University of Lyon as part of the Programme Investissements d’Avenir (ANR-16-IDEX-0005 to D. N.) and Higher Education Institutional Excellence Programme of the Ministry of Human Capacities in Hungary, within the framework of the Neurology thematic program of Semmelweis University (to K. F., E. K., B. Sz., J. R.).

## Supporting information

**S1 Table. Social distance in intentional conditions. Descriptive statistics and group differences**.

ASD: Autism Spectrum Disorder, NTP: Neurotypical Participant, N: sample size, SD: standard deviation

**S2 Table. HRV in explicit conditions. Descriptive statistics and group differences**. ASD: Autism Spectrum Disorder, NTP: Neurotypical Participant, N: sample size, SD: standard deviation

**S1 File. Questionnaires**

## Notes

### Competing Interest Statement

The authors have declared no competing interest.

### Author Declarations

Regional and Institutional Committee of Science and Research Ethics, Semmelweis University, Budapest, Hungary (SERKEB No.: 145/2019)

