## Supplement method for "Altered interpersonal distance regulation in autism spectrum disorder"

**Questionnaires**

**1.** **Autism-Spectrum Quotient (AQ).** To assess autistic traits in our sample, we used the AQ50 (Simon Baron-Cohen et al., 2001). The questionnaire consists of 50 questions about e.g. social skills, communication, or attention to details. Participants had to rate the items on a four-point Likert scale (1-4, indicating “definitely disagree”, “slightly disagree”, “slightly agree”, and “definitely agree”, respectively). When scoring the questionnaire, these options were dichotomized, all agreed answers were scored as 1, the other two options as 0 in non-reversed items, the opposite in the case of reverse items. Thus, the maximum score of the questionnaire was 50 (the clinical threshold for ASD >=32). (Chronbach’s alpha in our study = 0.92)

**2. Mentalization Questionnaire (MZQ).** Mentalization abilities were measured by the MZQ (Hausberg et al., 2012), which is a 15-item self-report questionnaire. MZQ has four subscales: ‘Refusing self-reflection’, ‘Emotional awareness’, ‘Psychic equivalence mode’ and ‘Regulation of affect’, rating on a five-point scale, (from “strongly disagree” to “strongly agree”). The total score ranges from 15 to 75 (indicating from good to poor mentalization skills, respectively). According to Hausberg et al. (2012), (n=434), the Cronbach’s alpha of the MZQ total score is 0.81, which can be considered as good consistency, but internal consistencies for the subscales ranged from 0.54 (Regulation of affect), to 0.72 (Psychic equivalence mode) among neurotypical participants which have been considered insufficient to satisfactory according to the COTAN criteria (Paridaens, 2016). For this reason, we use the total score in this study. (Chronbach’s alpha = 0.81)

**3. Adult ADHD Self-Report Scale v1.1 (ASRS).** The ASRS is a questionnaire that evaluates ADHD symptoms as defined in the Diagnostic and Statistical Manual of Mental Disorders fourth edition (DSM-IV) (Silverstein et al., 2019). ASRS is divided into two parts: Part A (consisting of 6 questions) and Part B (12 questions). For each item, respondents are asked to indicate how often the stated symptom occurred over the prior six months, with five options: never, rarely, sometimes, often, or very often. Part B items provide insight into the frequency of symptoms and can be used to help elicit other symptoms the patient may suffer from. According to Silverstein et al. (2019), the reliability of the ASRS and subscales is high (Cronbach’s α >= 0.84). In our study Cronbach’s alpha (total) = 0.87, part A = 0.61, part B = 0.85)

**4. Adult Attachment Scale (AAS).** We used the Dependence and Anxiety subscales of the AAS (Collins, 1996) to measure the attachment style of our participants. The questionnaire consists of 18 items, with three subscales. Participants rated on a scale from one (“Not typical to me at all”) to five (“Very typical to me”) how they think they function/would function in a romantic relationship. Collins et al. reported Cronbach's alpha coefficients of 0.69 for close, 0.75 for depend, and 0.72 for anxiety subscale (Collins & Read, 1990). Later authors found a slightly modified structure more useful and consistent. The scale measures adult attachment styles named *secure* (high scores on close and depend subscales, low score on anxiety subscale), *anxious* (high score on anxiety subscale, moderate scores on close and depend subscales), and *avoidant* (low scores on close, depend, and anxiety subscales) (Collins, 1996). We decided to use two subscales (anxious and avoidant) of attachment style in this study (Chronbach’s alpha total = 0.88, anxious = 0.85, avoidant = 0.85).

**5. State-Trait Anxiety Inventory (STAI).** STAI is a commonly used measure of trait and state anxiety (Spielberger et al., 1983). It contains 20 items for assessing state and trait anxiety. All items are rated on a four-point scale (from “Almost Never” to “Almost Always”). Higher scores indicate greater anxiety. Internal consistency coefficients for the scale have ranged from .86 to .95; test-retest reliability coefficients have ranged from .65 to .75 over a 2-month interval (Spielberger et al., 1983). In our study, we measured only trait anxiety. Chronbach’s alpha = 0.92).
