## Supplement Table1 for "Altered interpersonal distance regulation in autism spectrum disorder"

**S1 Table. Social distance in intentional conditions. Descriptive statistics and group differences.**

ASD: Autism Spectrum Disorder, NTP: Neurotypical Participant, N: sample size, SD: standard deviation

| Condition | | | | Mean *(SD)* | | | Mann-Whitney  U *(W)* | *p* | *Rank biserial correlation* |
| --- | --- | --- | --- | --- | --- | --- | --- | --- | --- |
|  |  |  |  | **NTP** *(N=21)* | | **ASD** *(N=22)* |  |  |  |
| Eye | Active | Self | 69.667 (33.95) | | 123.227 (86.42) | | 106.500 | **0.003** | -0.539 |
|  |  | Other | 73.000 (31.23) | | 111.455 (40.46) | | 101.000 | **0.002** | -0.563 |
|  | Passive | Self | 61.810 (42.80) | | 100.955 (65.78) | | 128.000 | **0.013** | -0.444 |
|  |  | Other | 67.238 (31.22) | | 98.909 (42.53) | | 116.000 | **0.005** | -0.498 |
| No-eye | Active | Self | 66.571 (31.52) | | 95.682 (54.69) | | 152.000 | 0.056 | -0.342 |
|  |  | Other | 75.762 (33.21) | | 98.182 (44.64) | | 169.000 | 0.127 | -0.273 |
|  | Passive | Self | 55.857 (23.56) | | 101.636 (58.01) | | 96.000 | **0.001** | -0.584 |
|  |  | Other | 71.619 (27.62) | | 99.318 (46.08) | | 142.000 | **0.032** | -0.383 |
