## Supplement Table2 for "Altered interpersonal distance regulation in autism spectrum disorder"

**S2 Table. HRV in explicit conditions. Descriptive statistics and group differences.**

ASD: Autism Spectrum Disorder, NTP: Neurotypical Participant, N: sample size, SD: standard deviation

| Condition | | | Mean *(SD)* | | Mann-Whitney  U *(W)* | *p* | *Rank biserial correlation* |
| --- | --- | --- | --- | --- | --- | --- | --- |
|  |  |  | **NTP** *(N=20)* | **ASD** *(N=19)* |  |  |  |
| Eye | Active | Self | 30.186 (21.72) | 26.330 (25.54) | 229.000 | 0.283 | 0.205 |
|  |  | Other | 30.865 (24.80) | 24.695 (22.04) | 229.000 | 0.283 | 0.205 |
|  | Passive | Self | 33.522 (24.86) | 28.227 (26.85) | 234.000 | 0.224 | 0.231 |
|  |  | Other | 36.623 (37.45) | 20.639 (14.61) | 253.000 | **0.034** | 0.402 |
| No-eye | Active | Self | 28.689 (15.04) | 24.830 (13.46) | 213.000 | 0.531 | 0.121 |
|  |  | Other | 29.722 (15.41) | 26.520 (16.55) | 205.000 | 0.478 | 0.139 |
|  | Passive | Self | 34.884 (22.07) | 24.777 (21.89) | 234.000 | 0.119 | 0.300 |
|  |  | Other | 29.171 (13.76) | 21.162 (14.81) | 239.000 | **0.039** | 0.398 |
